# Distinguishing Admissions Specifically for COVID-19 from Incidental SARS-CoV-2 Admissions: A National Retrospective EHR Study

**DOI:** 10.1101/2022.02.10.22270728

**Authors:** Jeffrey G Klann, Zachary H Strasser, Meghan R Hutch, Chris J Kennedy, Jayson S Marwaha, Michele Morris, Malarkodi Jebathilagam Samayamuthu, Ashley C Pfaff, Hossein Estiri, Andrew M South, Griffin M Weber, William Yuan, Paul Avillach, Kavishwar B Wagholikar, Yuan Luo, The Consortium for Clinical Characterization of COVID-19 by EHR (4CE), Gilbert S Omenn, Shyam Visweswaran, John H Holmes, Zongqi Xia, Gabriel A Brat, Shawn N Murphy

## Abstract

**Background:** Admissions are generally classified as COVID-19 hospitalizations if the patient has a positive SARS-CoV-2 polymerase chain reaction (PCR) test. However, because 35% of SARS-CoV-2 infections are asymptomatic, patients admitted for unrelated indications with an incidentally positive test could be misclassified as a COVID-19 hospitalization. EHR-based studies have been unable to distinguish between a hospitalization specifically for COVID-19 versus an incidental SARS-CoV-2 hospitalization. Although the need to improve classification of COVID-19 disease vs. incidental SARS-CoV-2 is well understood, the magnitude of the problems has only been characterized in small, single-center studies. Furthermore, there have been no peer-reviewed studies evaluating methods for improving classification.

**Objective:** The aims of this study were to: first, quantify the frequency of incidental hospitalizations over the first fifteen months of the pandemic in multiple hospital systems in the United States; and second, to apply electronic phenotyping techniques to automatically improve COVID-19 hospitalization classification.

**Methods:** From a retrospective EHR-based cohort in four US healthcare systems in Massachusetts, Pennsylvania, and Illinois, a random sample of 1,123 SARS-CoV-2 PCR-positive patients hospitalized between 3/2020–8/2021 was manually chart-reviewed and classified as admitted-with-COVID-19 (incidental) vs. specifically admitted for COVID-19 (for-COVID-19). EHR-based phenotyping was used to find feature sets to filter out incidental admissions.

**Results:** EHR-based phenotyped feature sets filtered out incidental admissions, which occurred in an average of 26% of hospitalizations (although this varied widely over time, from 0%-75%). The top site-specific feature sets had 79-99% specificity with 62-75% sensitivity, while the best performing across-site feature set had 71-94% specificity with 69-81% sensitivity.

**Conclusions:** A large proportion of SARS-CoV-2 PCR-positive admissions were incidental. Straightforward EHR-based phenotypes differentiated admissions, which is important to assure accurate public health reporting and research.

## Introduction

Despite the ongoing COVID-19 pandemic and the dozens of research groups and consortia around the world that continue to utilize clinical data available in Electronic Health Records (EHR), critical gaps remain in both our understanding of COVID-19 and how to accurately predict poor outcomes including hospitalization and mortality.[1–4]

One of the most prominent gaps in the field is how to distinguish hospital admissions specifically for COVID-19-related indications (e.g., severe disease with respiratory failure) from an incidentally positive SARS-CoV-2 PCR test in admissions for an unrelated reason (e.g., broken leg). Approximately 800,000 new SARS-CoV-2 cases are being reported daily, and approximately 150,000 patients are hospitalized with a positive SARS-CoV-2 PCR test.[5] Misclassification of incidental COVID-19 during hospitalizations is common[5] and raises research and public health concerns. For example, deleterious effects on healthcare system resource disbursement or utilization as well as on local and regional social and economic structure and function can result from inaccurate reporting of incidental cases of SARS-CoV-2.

Misclassification in research studies occurs because patients are usually considered COVID-19 patients if they have a recent positive SARS-CoV-2 PCR test or the ICD-10 diagnosis code U07, which, according to guidelines, is equivalent to a positive test.[6] This approach has been used in most COVID-19 studies published to date[7,8] and is in line with CDC guidelines, which treat positive SARS-CoV-2 PCR tests as confirmed cases.[9] Given that at least 35% of SARS-CoV-2 cases are asymptomatic, patients seeking unrelated care are erroneously classified as COVID hospitalizations.[10–14] The magnitude of this misclassification has increased over time as healthcare systems began to be less restrictive after the second wave and elective surgeries were again performed starting in the second quarter of 2021.

A potential solution is EHR-based phenotyping, which identifies patient populations of interest based on proxies derived from EHR observations. EHR phenotypes are developed by first performing manual chart review to classify cases and then applying a machine learning or statistical reasoning method to the EHR data to create an explainable predictive model.[15,16] For example, a phenotyping study of bipolar disorder found that true bipolar disorder correlated with a set of several EHR features.[17] Our previous work validated a “severe COVID-19” phenotype in the Consortium for Clinical Characterization of COVID-19 by EHR (4CE) network using both chart review and comparison across sites.[18,19] 4CE is a diverse international network oof over 300 hospitals engaged in collaborative COVID-19 research.[2,20,21]

The Massachusetts Department of Public Health has recently begun using a simple phenotype to report COVID-19 hospitalizations.[22,23] Although it is based on treatment recommendations and not a gold standard, it illustrates the interest in EHR-based phenotyping for COVID-19.

In this study, we utilized EHR data from 60 hospitals across four US healthcare systems in 4CE combined with clinical expertise, data analytics, and manual EHR chart review to determine whether patients admitted to the hospital and who had a positive SARS-CoV-2 PCR test were hospitalized for COVID-19 (for-COVID) or were admitted for a different indication and simply had an incidental positive test (incidental).

## Methods

We selected a sample of our 4CE sites across the US to participate in the development of our “for-COVID-19” hospitalization phenotype. These sites included Beth Israel Deaconess Medical Center (BIDMC), Mass General Brigham (MGB), Northwestern University (NWU), and University of Pittsburgh / University of Pittsburgh Medical Center (UPITT). Each site involved at least one *clinical* expert (for chart review and manual annotation) and one *data analytics* expert (to apply various analytic filtering approaches). Eligible patients for this study were those included in the 4CE COVID-19 cohort: all hospitalized patients (pediatric and adult) with their first positive SARS-CoV-2 PCR test seven days before to 14 days after hospitalization.[2]

### Chart Review

Each development site randomly sampled an equal number of admissions in each quarter (BIDMC, MGB) or month (NWU, UPITT) from their cohort of SARS-CoV-2 PCR positive patients over the period March 2020 until at least March 2021. Clinical experts reviewed the charts in the EHR and recorded whether these patients were admitted for COVID-19 related reasons as defined below. Participating sites and number of chart reviews are listed in Table 1.

**Table 1.** Participating healthcare systems’ overall characteristics and number and time period of chart reviews performed for this study.

| Participating Site | Hospitals | Inpatient Discharges Per Year | Number of Chart Reviews Performed | Chart Review Time Period<br>Start Date<br>End Date |
| --- | --- | --- | --- | --- |
| Beth Israel Deaconess Medical Center (BIDMC) | 1 | 40,752 | 400 | 3/2020<br>3/2021 |
| Mass General Brigham (MGB) | 10 | 163,521 | 406 | 3/2020<br>7/2021 |
| Northwestern University (NWU) | 10 | 103,279 | 70 | 3/2020<br>2/2021 |
| University of Pittsburgh / University of Pittsburgh Medical Center (UPITT) | 39 | 369,300 | 247 | 4/2020<br>8/2021 |

To develop chart review criteria, a 4CE sub-group met during March-July 2021. The group consists of about 20 researchers in 4CE, with a mixture of physicians, medical informaticians, and data scientists. In the process, dozens of real patient charts were considered, and edge cases were discussed until consensus was reached on the minimal chart review necessary to determine the reason a patient was hospitalized.

Based on the developed criteria (Table 2), chart reviewers (one per site, except at BIDMC where there were two) classified the patients based on review of primarily the admission note, discharge summary (or death note), and laboratory values for the hospitalization. Each site had IRB approval to view the charts locally and only de-identified aggregate summaries were presented to the sub-group. Each site summarized the chart reviews in a spreadsheet that was then linked to the site’s 4CE EHR data, wherein medical record numbers were replaced with 4CE’s patient pseudo-identifiers, and criteria classifications were coded as an integer. The chart review process is presented visually in (Figure 1).

**Table 2.** Summary of the chart-review criteria developed by the 4CE subgroup of physicians, medical informaticians, and data scientists.

| Chart Reviewed Classification | Criteria |
| --- | --- |
| Admitted Specifically for COVID-19 | Symptoms on admission **were attributable** to COVID-19 and clinicians admitted patients for COVID-19-related care. This includes:<br>- Respiratory insufficiency<br>- Blood clot to vital organs<br>- Hemodynamic changes<br>- Other common viral symptoms such as cough, fever, etc.<br>- Admitted for non-COVID-19 issue, but developed one of the above criteria while hospitalized |
| Admitted Incidentally with COVID-19 | Admission history was **unlikely** to be related to COVID-19 and clinicians did not specifically admit the patient for COVID-19-related care. This admission could be due to:<br>- Trauma<br>- Procedure or operation requiring hospitalization<br>- Term labor<br>- Alternative causes, including drug overdose, cancer progression, non-respiratory severe infection, etc. |
| Uncertain | Symptoms on admission **may have been** related to COVID-19 and clinicians considered COVID-19 exacerbation during hospitalization. This includes:<br>- Preterm labor<br>- Liver dysfunction<br>- Graft failure<br>- Immune system dysfunction<br>- Alternative causes including sickle cell crisis, failure to thrive, altered mental status |

**Figure 1.**
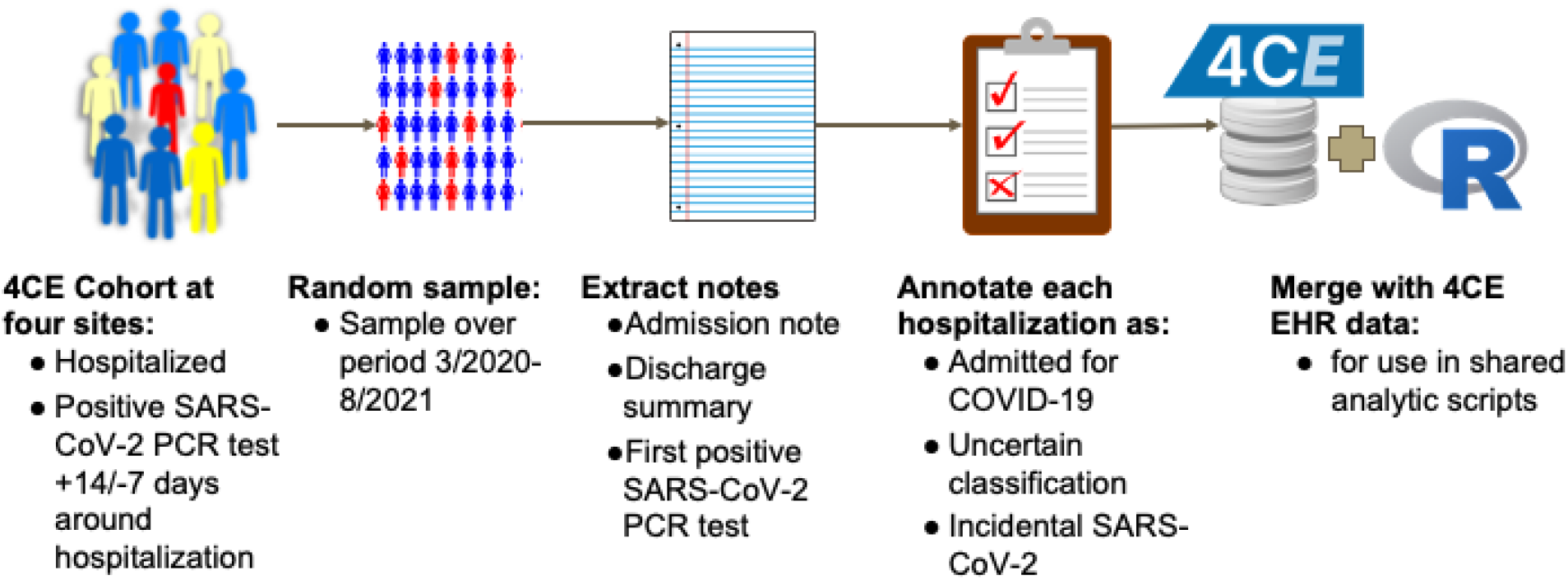
The chart-review process. At each site, an equal number of patients admitted with a positive SARS-CoV-2 PCR test were sampled by quarter or by month. A chart reviewer at the site examined primarily the admission note, discharge summary (or death note), and laboratory values for the hospitalization to classify as admitted for COVID-19, incidental SARS-CoV2, or uncertain. These classifications were then merged with 4CE EHR data for use with shared analytic scripts in R.

We developed an R script at MGB to perform basic data summarization. It did the following: calculated chart review summary statistics; aggregated data on ICD-10 diagnosis codes used during the hospitalization to compare to the chart review classification; generated a bubble plot that visualizes the change in proportion of hospitalizations specifically for COVID-19 among all chart reviews over the course of the pandemic, by month. A trendline was fitted with loess regression using ggplot2 and was weighted by the number of chart reviews performed that month. Each participating healthcare site ran the R script on their chart reviewed patient cohort.

### Phenotypes Using Hospital System Dynamics Phenotyping

We developed an algorithm as an R script to choose phenotypes of admissions specifically for COVID-19, using established hospital dynamics measures of ordering/charting patterns in the EHR (e.g., presence of laboratory tests rather than laboratory results).[16,24] The algorithm uses a variation of an Apriori itemset-mining algorithm.[25,26] Apriori, which has been employed in other EHR studies, employs a hill-climbing approach to find iteratively larger item sets that meet some summary statistic constraint.[27,28] The original algorithm chose rules that maximized positive predictive value (PPV) and had at least a minimum prevalence in the dataset. More recent variants use other summary statistics,[29] because PPV, which measures the likelihood a positive is a true positive, is highly affected by population prevalence (which shifts dramatically over time with COVID-19). Therefore, our algorithm used sensitivity and specificity. A visual representation of our algorithm is shown in (Figure 2). Itemsets of size 1 are chosen that meet certain minimum prediction thresholds, then these are combined into itemsets of size 2 and again filtered by the thresholds, and so forth up to a maximum itemset size.

**Figure 2.**
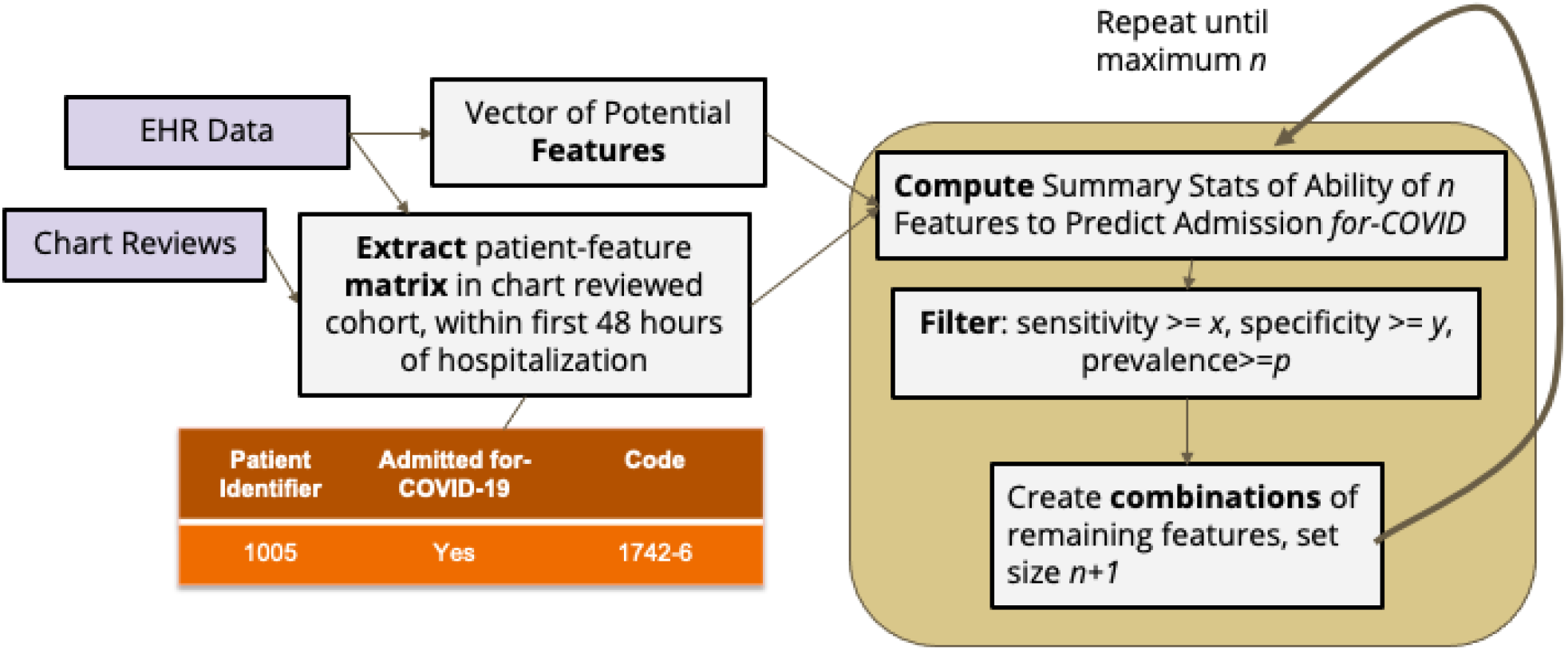
Design of the phenotyping algorithm. Predictive feature sets of iteratively larger size are selected based on their sensitivity and specificity in correctly identifying COVID-19-specific admissions using 4CE EHR data and chart reviews. We chose the following parameters after testing various thresholds at all four sites: AND feature sets, x=0.40, y=0.20, p=0.30; OR feature sets x=0.10, y=0.50, p=0.20; single features: x=y=p=0

We applied our algorithm to find patterns in 4CE EHR data at each site using presence of medications, laboratory tests, and diagnoses to select the best phenotypes. We further compared the output at each site to see if there were similarities, e.g., transfer-learning was applicable. We considered two cases: data that would be available in near-real time during a hospitalization (laboratory tests) and data that would be available for a retrospective analysis (including laboratory and medication facts and diagnosis codes - which are usually not coded until after discharge).

Sites exported phenotypes with sensitivity of at least 0.60, ordered by specificity in descending order. (Site B applied a slightly lower sensitivity threshold because no phenotypes with sensitivity of at least 0.60 were available.) Specificity was chosen as the sorting variable because it measures the phenotype’s ability to detect and remove incidental SARS-CoV-2 admissions—a good measure of overall performance. Sensitivity, on the other hand, measures the ability to select for-COVID admissions, which can be easily maximized by simply selecting all patients. Groups of phenotypes were manually summarized into conjunctive normal form by combining AND and OR phenotypes at each site when possible and reporting a sensitivity and specificity range for the final combined phenotype. We excluded feature sets that were more complex but with the same performance as a simpler feature set.

We also ran our phenotyping program to find the most predictive single items at each site during every 6-month period of the pandemic, beginning January 2020. This analysis allowed us to examine the trend of Hospital System Dynamics (HSD) as the pandemic progressed.

The final piece of analysis involved selecting multi-site phenotypes and plotting their performance over time. First, we isolated the components of phenotypes that appeared at multiple sites, resulting in three multisite feature sets. We manually added/removed OR components based on performance at MGB (because adding too many OR components degrades the specificity). We ran these constructed phenotypes at each site to ascertain their performance characteristics.

### Temporal Visualization of Phenotypes

We also developed a visualization used at each site. The visualization shows three lines: a solid line shows the total number of patients in the site’s 4CE cohort (i.e., admitted with a positive SARS-CoV-2 PCR test); a dashed line shows the total number of those patients after filtering to select patients admitted specifically for COVID-19 (i.e., removing all patients that do not meet the phenotyping feature set criteria); a dotted line shows the difference of the solid line and the dashed line (i.e., patients removed from the cohort in the dashed line). Dots on the graph visualize the performance on the chart reviewed cohort. Green dots on each line show patients that were correctly classified by the phenotype, according to the chart review. Likewise, orange dots on each line show incorrect classifications. Dot size is proportional to the number of chart reviews.

Importantly, all review and analysis were performed by local experts at each site, and only the final aggregated results were submitted to a central location for finalization. This approach is one of the hallmarks of 4CE - keeping data close to local experts and only sharing aggregated results. It reduces regulatory complexity around data sharing and keeps those who know the data best involved in the analysis.

All our software tools were implemented as R programs. They were developed at MGB and tested by all four sites. The code is available as open source.[30]

Institutional Review Board Approval was obtained at Beth Israel Deaconess Medical Center, Mass General Brigham, Northwestern University, and University of Pittsburgh. Participant informed consent was waived by each IRB because the study involved only retrospective data and no individually identifiable data was share outside of each site’s local study team. Site names were anonymized (to sites A, B, C, and D) to comply with hospital privacy policies. At MGB and BIDMC, any counts of patients were blurred with a random number +/- 3 before being shared centrally. Our previous work shows that, for large counts, pooling blurred counts has minimal impact on the overall accuracy of the statistics.[31] At all sites, any counts with fewer than three were censored. All other statistics (e.g., percentages, differences, confidence intervals, p-values) were preserved.

## Results

### Chart Review

The final chart review criteria are shown in (Table 2). (See Methods for details.) Across the four sites, 68% of patients were admitted *for* COVID-19, 26% of patients were admitted with *incidental* SARS-CoV-2, and 6% were uncertain (Table 3). The four sites included Beth Israel Deaconess Medical Center (BIDMC), Mass General Brigham (MGB), University of Pittsburgh / University of Pittsburgh Medical Center (UPITT), and Northwestern University (NWU). A site-by-site breakdown both overall and by individual criteria is also shown in (Table 3). Plots of the proportion of hospitalizations specifically for COVID-19 among all chart reviews by month over the course of the pandemic are shown in (Figure 3). Finally, (Table 4) shows the top-10 ICD-10 diagnoses that were assigned to patients with a date in the first 48 hours after admission in *for-* COVID vs. *incidental* COVID patients (Table 4). In all results, the sites are labeled with a random but consistent letter (A, B, C, or D) to comply with hospital privacy policies.

**Table 3.** Proportion of chart-reviewed patients admitted specifically for COVID-19 vs. admitted with incidental SARS-CoV-2, overall and stratified by site, with a detailed criteria breakdown. A detailed breakdown at Site D could not be included because their process did not record the specific criteria for each classification.

| COVID Classification | Category | Site A | Site B | Site C | Site D | Overall |
| --- | --- | --- | --- | --- | --- | --- |
| **Admitted Specifically for COVID-19** | *ALL* | 71% | 84% | 73% | 60% | 68% |
|  | Respiratory insufficiency | 50% | 51% | 52% | - |  |
|  | Blood clot | 1% | 3% | 0% | - |  |
|  | Hemodynamic | <1% | 0% | 0% | - |  |
|  | Other symptomatic COVID-19 | 18% | 27% | 20% | - |  |
|  | Not admitted for COVID-19 but developed one of the above criteria | 2% | 3% | 2% | - |  |
| **Admitted Incidentally with COVID-19** | *ALL* | 21% | 13% | 22% | 36% | 26% |
|  | Trauma | 0% | 0% | 0% | - |  |
|  | Procedure | 2% | 0% | 4% | - |  |
|  | Full-term labor | 4% | 4% | 0% | - |  |
|  | Other not COVID-19 | 13% | 9% | 18% | - |  |
| **Uncertain** | *ALL* | 8% | 3% | 4% | 4% | 6% |
|  | Other possible COVID-19 | 8% | 3% | 4% | - |  |
|  | Early labor | <1% | 0% | 0% | - |  |
|  | Liver dysfunction | <1% | 0% | 0% | - |  |
|  | Graft failure | 0% | 0% | 0% | - |  |
|  | Immune dysfunction | <1% | 0% | 0% | - |  |

**Table 4.** Top ten ICD-10 diagnoses among patients chart reviewed as admitted specifically for COVID-19 and those admitted with incidental COVID-19, with the proportion of patients at each site. Each patient might have multiple diagnoses, and therefore the sum might be greater than 100%.

| ICD-10 Diagnosis | Site A | Site B | Site C | Site D |
| --- | --- | --- | --- | --- |
| **Admitted specifically for COVID-19** |  |  |  |  |
| U07.1 Covid-19 | 92% | 92% | 80% | 95% |
| J12.89 Other Viral Pneumonia | 44% | 41% | 35% | 70% |
| I10 Essential (Primary) Hypertension | 39% | 27% | 41% | 37% |
| J96.01 Acute Respiratory Failure With Hypoxia | 26% | 34% | 31% | 58% |
| E78.5 Hyperlipidemia, Unspecified | 28% | 7% | 38% | 46% |
| N17.9 Acute Kidney Failure, Unspecified | 25% | 7% | 22% | 39% |
| K21.9 Gastro-Esophageal Reflux Disease Without Esophagitis | 22% | 2% | 31% | 26% |
| Z87.891 Personal History of Nicotine Dependence | 18% | 2% | 24% | 27% |
| R09.02 Hypoxemia | 29% | 25% | 12% | 17% |
| J12.82 Pneumonia due to COVID-19 | 25% | 20% | 22% | 15% |
| **Admitted Incidentally with COVID-19** |  |  |  |  |
| U07.1 Covid-19 | 74% | 56% | 73% | 85% |
| N17.9 Acute Kidney Failure, Unspecified | 14% | 11% | 22% | 17% |
| E11.22 Type 2 Diabetes Mellitus with Diabetic Chronic Kidney Disease | 6% | 11% | 13% | 15% |
| E11.9 Type 2 Diabetes Mellitus Without Complications | 11% | 11% | 7% | 11% |
| D64.9 Anemia, Unspecified | 19% | 11% | 9% | 6% |
| E87.2 Acidosis | 6% | 11% | 5% | 10% |
| J12.89 Other Viral Pneumonia | 2% | 22% | 7% | 12% |
| J96.01 Acute Respiratory Failure With Hypoxia | 8% | 11% | 7% | 8% |
| D69.6 Thrombocytopenia, Unspecified | 7% | 11% | 11% | 7% |
| N18.6 End Stage Renal Disease | 7% | 11% | 9% | 5% |

**Figure 3.**
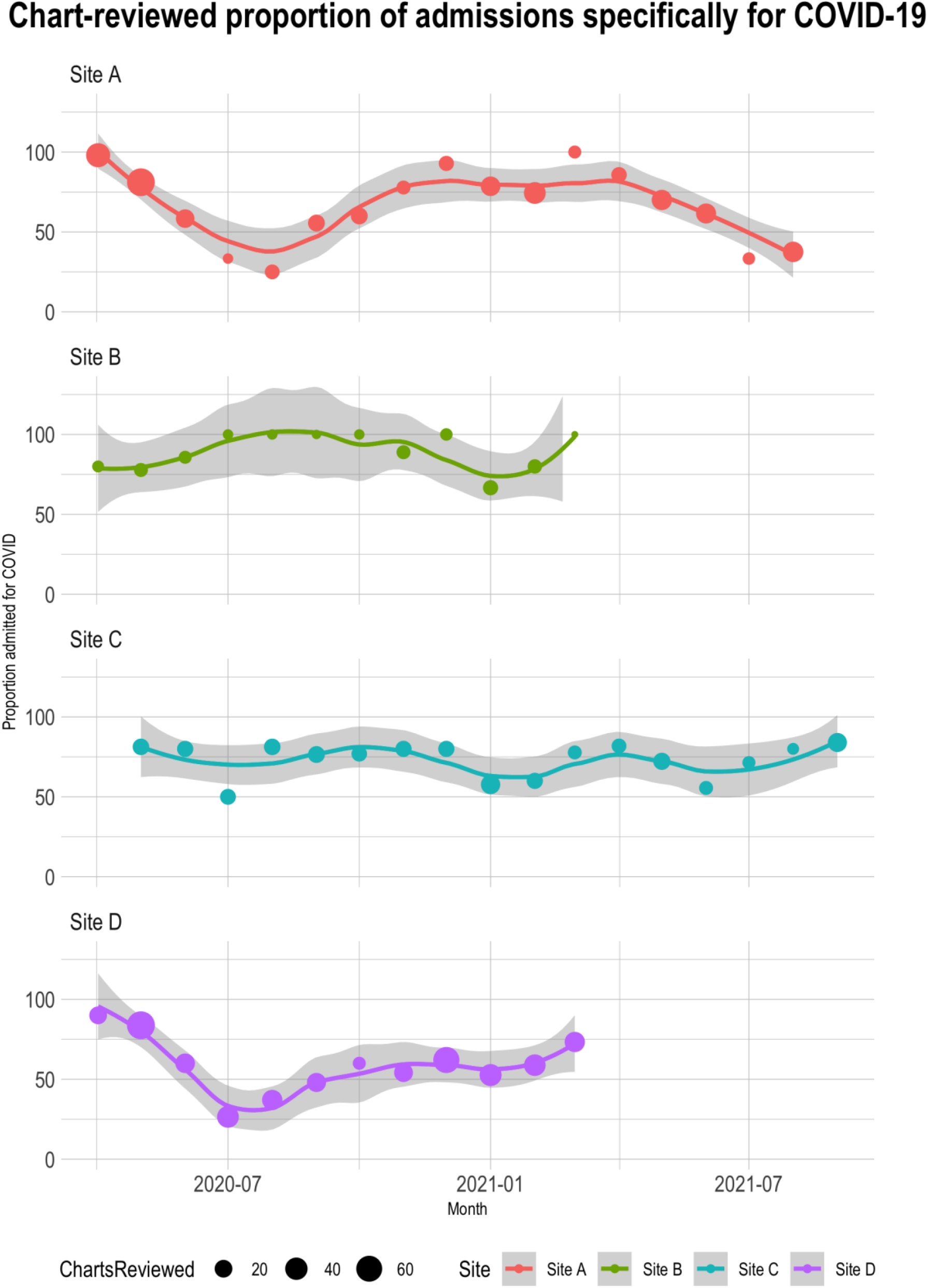
Chart-reviewed proportion of admissions specifically for COVID-19. The proportion of hospitalizations specifically for COVID-19 among all chart reviews by month at each site. Bubble size shows the relative number of patient chart reviews performed that month. The trendline was weighted by bubble size and was performed using loess regression. Note that the y-axis and confidence interval limits extend above 100%.

### Phenotypes Using Hospital System Dynamics

Each site ran our Hospital System Dynamics (HSD) program to choose phenotypes of patients admitted *for-COVID* vs. patients admitted *incidentally with COVID*. The input of the program includes the chart-reviewed classifications and patient-level EHR data on the presence of laboratory tests, medications, and diagnosis codes that are dated within 48 hours of admission. (Table 5) shows the top feature sets generated by the program at each site both for phenotypes that use data that could be available immediately (“real time”) and phenotypes using all data available after discharge (“retrospective”). We also report prevalence at each site among all SARS-CoV-2 PCR positive hospitalizations (not just among chart-reviewed patients), which is the proportion of patients meeting the criteria of the feature sets.

**Table 5.** Top phenotyping feature sets by specificity, with a sensitivity of at least 0.60 for detecting admissions specifically for COVID-19. The table is grouped into feature sets involving potentially real-time data (laboratory tests) and all available data (presence of laboratory tests, medications, and diagnosis codes). Ranges are shown in the summary statistics because multiple rules with similar performance were summarized using conjunctive normal form.

| Phenotyping Feature Set | Site | Sensitivity | Specificity | Prevalence |
| --- | --- | --- | --- | --- |
| **“Real-time” phenotypes (labs only)** |  |  |  |  |
| CRP AND<br>(total bilirubin OR ferritin OR LDH) AND<br>(lymphocyte count OR neutrophil count) AND<br>cardiac troponin | Site D | 0.65-0.72 | 0.85 | 67-71% |
| Ferritin AND LDH AND cardiac troponin AND (INR<br>OR PTT OR lymphocyte count OR neutrophil<br>count) | Site D | 0.62-0.69 | 0.85 | 67-71% |
| CRP AND (LDH AND/OR Ferritin) AND cardiac<br>troponin | Site A | 0.67-0.70 | 0.89-0.90 | 72-77% |
| Procalcitonin OR d-dimer OR CRP OR<br>cardiac troponin OR ferritin | Site A | 0.63-0.87 | 0.73-0.85 | 65-85% |
| Any two of: procalcitonin, LDH, CRP | Site B | 0.56-0.58 | 0.67 | 63-67% |
| D-dimer OR ferritin OR CRP | Site C | 0.26-0.37 | 0.86-0.93 | 54-58% |
| **“Retrospective” phenotypes (labs, meds, and diagnosis codes)** |  |  |  |  |
| Total bilirubin AND<br>(ferritin OR LDH OR lymphocyte count OR<br>neutrophil count) AND diagnosis of Other Viral<br>Pneumonia (J12.89) | Site D | 0.62-0.64 | 0.92 | 46-48% |
| Diagnosis of: Other Viral Pneumonia (J12.89) OR<br>Acute Respiratory Failure with Hypoxia (J96.01)<br>OR Anemia (D64.9) | Site D | 0.70-0.74 | 0.82-0.88 | 50-63% |
| Diagnosis of: Other Viral Pneumonia (J12.89) OR<br>Supplemental Oxygen (severe) | Site D | 0.75 | 0.82 | 61% |
| CRP AND (LDH OR ferritin)<br>AND cardiac troponin | Site A | 0.70 | 0.89 | 74-77% |
| Remdesivir OR procalcitonin OR Other Viral<br>Pneumonia (J12.89) OR Nonspecific Abnormal<br>Lung Finding (R91.8) OR Shortness of Breath<br>(R06.02) OR Other COVID Disease (J12.82) | Site A | 0.68-0.72 | 0.85-0.95 | 58-74% |
| Hypoxemia (R09.02) OR other Coronavirus as<br>Cause of Disease (B97.29) OR Shortness of<br>Breath (R06.02) OR Pneumonia (unspecified<br>organism) (J18.9) OR acute respiratory failure<br>with hypoxia (J96.01) OR Nonspecific Abnormal<br>Lung Finding (R91.8) | Site B | 0.63-0.68 | 0.89-0.99 | 54-67% |
| D-dimer OR ferritin OR CRP OR Other Viral<br>Pneumonia (J12.89) OR acute respiratory failure<br>with hypoxia (J96.01) | Site C | 0.71-0.75 | 0.79-0.86 | 52-58% |

We examined the top individual EHR data elements over time at all sites. In the first half of 2020, a diagnosis of “other viral pneumonia” (J12.89) was the only strong predictor of an admission specifically for COVID-19 across all four sites. In the second half of 2020, the phenotyping algorithm began selecting laboratory tests, including CRP, troponin, ferritin, and LDH. Also, the diagnosis “other COVID disease” (B97.29) began to be used at Site B. By 2021, remdesivir and the diagnosis “pneumonia due to COVID-19” (J12.82) additionally came into widespread use and became predictive of admissions specifically for COVID at Site A.

### Temporal Visualization of Phenotypes

The three multisite phenotypes (derived from common elements in Table 5) and their performance are shown in (Table 6), with the top rule at each site in bold type. In (Figure 4), we plotted the performance of the top phenotype at each site (the boldfaced rows in Table 6) using the temporal phenotype visualization described in the Methods. (The top phenotype involved all data types at every site except Site C, where diagnoses alone performed better.)

**Table 6.** The best multisite phenotyping feature sets and their overall performance characteristics. The top-performing phenotype at each site is boldfaced. The multisite phenotypes were derived from (Table 5), by selecting components of phenotypes that appeared at multiple sites.

| Phenotyping Feature Set | Description | Sensitivity, Specificity |
| --- | --- | --- |
| Other Viral Pneumonia OR Acute Respiratory Failure with Hypoxia OR Shortness of Breath OR Abnormal Lung Finding | Diagnoses mentioned in top feature sets at >1 site | Site A: 0.79,0.72<br>Site B: 0.88, 0.85<br>**Site C: 0.69,0.90**<br>Site D: 0.64,0.58 |
| CRP AND Ferritin | Labs mentioned in top feature sets at all four sites | Site A: 0.76,0.85<br>Site B: 0.88, 0.85<br>Site C: 0.42, 0.98<br>Site D: 0.66, 0.55 |
| Remdesivir OR Oxygen (severe) OR Dx of Other Viral Pneumonia | All items mentioned at multiple sites in OR feature sets | **Site A: 0.74,0.91**<br>**Site B: 0.81, 0.94**<br>Site C: 0.60,0.92<br>**Site D: 0.61,0.71** |

**Figure 4.**
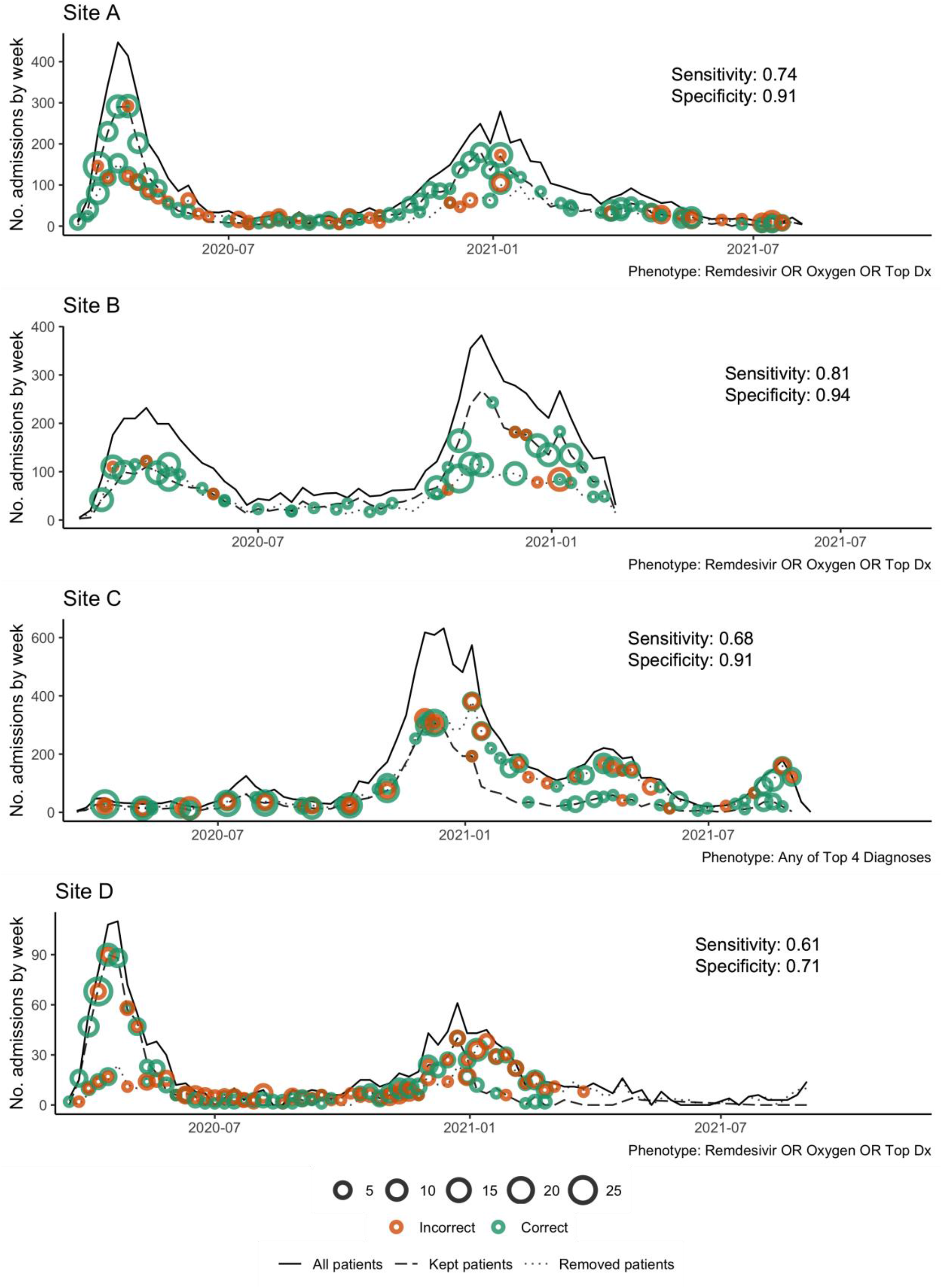
Phenotype performance over time at each site. Performance of the top phenotyping feature set (Table 6) at each site. Y-axis is number of admissions per week, X-axis is week, and overall sensitivity and specificity shown on each figure panel. Solid lines show the total number of weekly admissions for patients with a positive SARS-CoV-2 PCR test. Dashed lines show the number of weekly admissions after filtering to select patients admitted specifically for COVID-19 (i.e., removing all patients that do not meet the phenotyping feature set criteria). Dotted line shows the difference of the solid line and the dashed line (i.e., patients removed from the cohort in the dashed line). Green dots indicate correct classification by the phenotype according to chart review. Orange dots indicate incorrect classification. Dot size is proportional to the number of chart reviews.

## Discussion

### Principal Results and Analysis

The COVID-19 pandemic has lasted for over two years, with multiple waves across the world. Although hospital systems have been cyclically overwhelmed by patients seeking care for COVID-19, as healthcare systems began to open up before the second wave, elective surgeries were again performed starting in the later part of 2020, and especially in the second quarter of 2021 many have approached the healthcare system for health issues (e.g., accidents, strokes, etc.) while incidentally infected with SARS-CoV-2.[32] This, along with the high false positive rate of SARS-CoV-2 PCR tests in some situations,[33–36] has led to increasing numbers of misclassified patients in analyses of COVID-19 characteristics and severity. This could be creating significant detection and reporting bias, leading to erroneous conclusions.[10–13] This study presents a multi-institutional characterization of 1,123 hospitalized patients either incidentally infected with SARS-CoV-2 or specifically hospitalized for COVID-19 in four healthcare systems across multiple waves using consensus-based chart review criteria.

We applied an itemset-mining approach and established Hospital System Dynamics (HSD) principles to phenotype SARS-CoV-2 PCR-positive patients who were admitted specifically for COVID-19, using data on charting patterns (e.g., presence of laboratory tests within 48 hours of admission) rather than results (e.g., laboratory results).[16,37] HSD examines healthcare process data about a hospitalization, such as ordering/charting patterns, rather than the full dataset. For example, to study severely ill patients, HSD might select patients with a high total number of labs for a patient on the day of admission. This could be an indirect measure of clinical suspicion of disease complexity or severity. Previous work shows that proxies such as total number of laboratory tests on the day of admission or the time of day of laboratory tests can be highly predictive of disease course.[24,37] Our methods sorted out who was treated for COVID-19 automatically, over time, with specificities above 0.70 even for some phenotypes discovered at a single site and applied to all four. We focused on specificity because the goal was to remove false positives (i.e., incidental SARS-CoV-2) from the cohort.

Our chart review protocol illustrates that patients who were admitted and have a positive SARS-CoV-2 PCR test were more likely to be admitted specifically for COVID-19 when disease prevalence was high (at least prior to Omicron). However, during periods in which healthcare systems were less restrictive (i.e., resumed routine surgeries), a secondary measure/phenotype is critical for accurately classifying admissions specifically for SARS-CoV-2 infection.

As expected, we observed a lower proportion of hospitalizations specifically for COVID-19 in the summer months when disease prevalence was lower (Figure 3). One would expect this, because there were fewer overall admissions as hospitals were recovering from the previous wave.

As expected, the top chart review criteria (Table 3) were *respiratory insufficiency* in admissions specifically for COVID-19 and *other* for incidental and uncertain admissions with SARS-CoV-2. Surprisingly, 10-20% of patients admitted with incidental SARS-CoV-2 were diagnosed with pneumonia, respiratory failure, or acute kidney injury (Table 4). This could reflect data collection issues, where some systems might repeat past problems automatically at hospital admission. In the case of codes for acute kidney injury, further investigation is needed to determine whether SARS-CoV-2-associated acute kidney injury (including COVID-19-associated nephropathy) occurs in patients we otherwise classified as having incidental admissions.[38]

Healthcare systems are beginning to explore phenotyping feature sets to report admissions specifically for COVID-19. Starting January 2022 in Massachusetts, hospitals began reporting the number of for-COVID hospitalizations as the count of admitted patients with both a SARS-CoV-2 positive test and a medication order for dexamethasone.[22,23] This simple phenotype was designed by the Massachusetts Department of Public Health as a first attempt, and it was based only on treatment recommendations for moderate-to-severe COVID-19 with hypoxia. It was not validated against a gold standard. Nonetheless, it illustrates the interest in EHR-based phenotyping for COVID-19.

Phenotypes with diagnosis codes tended to be the best performing predictors of admissions specifically for COVID-19. This could be because diagnosis codes represent either a clinically informed conclusion or a justification for ordering a test (implying the clinician suspected COVID-19). However, diagnoses are less prevalent in the population than laboratory tests and might not cover the entire population of admissions for COVID-19. Further, diagnoses early in hospitalization also do not always reflect the patient’s eventual diagnosis or hospital-related complications that are more accurately reflected in discharge diagnoses. There was also some heterogeneity in the diagnoses used at different sites (e.g., B97.29 “other COVID disease” was a top predictor only at Site B). In addition, presence of laboratory tests are useful for real-time detection systems because diagnosis codes usually are assigned after discharge. Clusters of tests for inflammatory markers (e.g., LDH, CRP, and ferritin) appeared across most sites as predictive of hospitalizations specifically for COVID-19, which fits intuitively because one of the underlying systemic pathophysiological mechanisms of SARS-CoV-2 is thought to be an inflammatory process,[39,40] and guidelines therefore have encouraged health care providers to check inflammatory markers on COVID-19 admissions.[41,42] Many of these inflammatory labs are not routinely ordered on all hospitalized patients and would therefore be expected to help distinguish COVID-19 from other patients. However, laboratory protocol differences across sites may have reduced generalizability for this metric.

Our methods generated pairs of items using OR and groups of up to 4 using AND logical operators. Our feature sets were somewhat vulnerable to the problem that specificity decreases when multiple elements are combined with OR although, in general, OR feature sets performed better across sites because they could be designed to choose the top performing elements at each site.

In addition to site differences, we also found changing disease management patterns over time. At the start of the pandemic, the only predictive phenotype was a pneumonia diagnosis. As standard COVID-19 order recommendations began to appear, laboratory orders became more consistent and predictive. Next, remdesivir began to be administered regularly. Finally, COVID-specific ICD-10 codes began to appear.

Overall, we found that an informatics-informed phenotyping approach successfully improved classification of for-COVID vs. incidental SARS-CoV-2 positive admissions, though generalizability was a challenge. Although some transfer learning is apparent (i.e., a few phenotypes performed well across sites), local practice and charting patterns reduced generalizability. Specifically, phenotypes involving only laboratory tests did not perform well at Site C, because the prevalence of these labs was low in the overall EHR data. This could be due to a data extraction or mapping issue in the underlying data warehouse. BIDMC had lower performance than other sites on the cross-site rules but not on the site-specific rules, perhaps highlighting less typical clinician treatment patterns.

### Limitations

Although the current data start at the beginning of the pandemic, they do not include the current Omicron wave nor very much of the Delta wave. We believe that the techniques introduced here (if not the phenotypes themselves) will be applicable to these variants, and we are planning future studies to validate this.

Our phenotypes demonstrated some transfer learning but not enough to create a single phenotype applicable to all sites. Technically, our system used machine learning at individual sites, but results were manually aggregated across sites. Emerging techniques for federated learning[43] might reduce the manual work required and increase the complexity of possible cross-site phenotype testing.

Finally, an inherent weakness of EHR-based research is that EHR data do not directly represent the state of the patient, because some observations are not recorded in structured data, and some entries in the EHR are made for non-clinical reasons (e.g., to justify the cost of a test or to ensure adequate reimbursement for services). This is common to all EHR research efforts, and we mitigated this limitation by developing chart-verified phenotypes.

### Conclusion

At four healthcare systems around the US over an 18-month period starting in March 2020, we developed and applied standardized chart review criteria to characterize the correct classification of a hospitalization specifically for COVID-19 as compared to incidental hospitalization of a patient with a positive SARS-CoV-2 test or ICD code. Then we applied HSD and frequent itemset mining to electronic phenotyping to generate phenotypes specific to hospitalizations for COVID-19, and we showed how patterns changed over the course of the pandemic. Application of this approach could improve public health reporting, healthcare system resource disbursement, and research conclusions.

## Data Availability

The electronic health record datasets analyzed during the current study cannot be made publicly available due to regulations for protecting patient privacy and confidentiality. These regulations also prevent the data from being made available upon request from the authors. Any questions about the dataset can be directed to the corresponding author.

https://github.com/jklann/jgk-i2b2tools/tree/master/4CE_utils

## Acknowledgements

We would like to additionally thank: Trang Le, PhD for her assistance in developing the temporal phenotype visualization R script; Karen Olson, PhD for her knowledge of the R programming language and insights about the topic of this study; Margaret Vella for her administrative coordination; and Isaac Kohane, MD, PhD for convening, leading, and guiding the 4CE Consortium.

## Author Contributions

JGK and SNM conceptualized and designed the study. ZHS, GAB, JGK, and SNM conceptualized and designed the chart review process. JGK, ZHS, MRH, CJK, JSM, MM, MJS, ACP, GMW, WY, YL, SV, ZX, and GAB contributed to data collection (of EHR data and/or chart reviews). JGK, ZHS, MRH, CJK, JSM, MM, MJS, ACP, AMS, GMW, WY, PA, KBW, GSO, SV, JHH, ZX, GAB and SNM contributed to data analysis or interpretation. JGK, ZHS, MRH, CJK, MM, HE, AMS, GMW, PA, KBW, YL, GSO, JHH, ZX, GAB, and SNM contributed to drafting and revision of the manuscript. All authors approved the final draft of the manuscript. SNM contributed to grant funding.

## Conflicts of Interest

The authors declare that they have no conflicts of interest.

### Other interests

JGK reports a consulting relationship with the i2b2-tranSMART Foundation through Invocate, Inc. CJK reports consulting for UC Berkeley, USC, UCSF. AMS reports funding from NIH/NIDDK R01DK127208; NIH/NHLBI R01HL146818 and institutional pilot awards from Wake Forest School of Medicine. GMW reports consulting for the i2b2-tranSMART Foundation. PA reports consulting for CCHMC and BCH. ZX has received research support from the National Institute of Health, the Department of Defense, and Octave Biosciences, and has served on the scientific advisory board for Genentech / Roche. SNM reports professional relationships with the Scientific Advisory Board for Boston University, Universidad de Puerto Rico, University of California at Los Angeles (UCLA), University of Massachusetts Medical School (UMMS), and The Kenner Family Research Fund.

## Data Sharing

All data analysis code developed for this study is available at https://github.com/jklann/jgk-i2b2tools/tree/master/4CE_utils under the Mozilla Public License v2 with healthcare disclaimer.

## Abbreviations

4CE: Consortium for Clinical Characterization of COVID-19 by HER
BIDMC: Beth Israel Deaconess Medical Center
CDC: Centers for Disease Control and Prevention
COVID-19 / COVID: Coronavirus Disease 2019
EHR: Electronic Health Record
HSD: Hospital System Dynamics
ICD-10: International Classification of Diseases, Tenth Revision
IRB: Institutional Review Board
MGB: Mass General Brigham
NWU: Northwestern University
PCR: Polymerase Chain Reaction
PPV: Positive Predictive Value
UPITT: University of Pittsburgh / University of Pittsburgh Medical Center
SARS-CoV-2: Severe Acute Respiratory Syndrome Coronavirus 2

