## Appendix for "Distinguishing Admissions Specifically for COVID-19 from Incidental SARS-CoV-2 Admissions: A National Retrospective EHR Study"

James R Aaron, Giuseppe Agapito, Adem Albayrak, Giuseppe Albi, Mario Alessiani, Anna Alloni, Danilo F Amendola, Li L.L.J Anthony, Bruce J Aronow, Fatima Ashraf, Andrew Atz, Paul Avillach, Paula S Azevedo, James Balshi, Brett K Beaulieu-Jones, Douglas S Bell, Antonio Bellasi, Riccardo Bellazzi, Vincent Benoit, Michele Beraghi, José Luis Bernal-Sobrino, Mélodie Bernaux, Romain Bey, Alvar Blanco-Martínez, Martin Boeker, Clara-Lea Bonzel, John Booth, Silvano Bosari, Florence T Bourgeois, Robert L Bradford, Gabriel A Brat, Stéphane Bréant, Carlos Tadeu Breda Neto, Nicholas W Brown, William A Bryant, Mauro Bucalo, Anita Burgun, Tianxi Cai, Mario Cannataro, Aldo Carmona, Charlotte Caucheteux, Julien Champ, Jin Chen, Krista Chen, Luca Chiovato, Lorenzo Chiudinelli, Kelly Cho, James J Cimino, Tiago K Colicchio, Sylvie Cormont, Sébastien Cossin, Jean B Craig, Juan Luis Cruz-Bermúdez, Jaime Cruz-Rojo, Arianna Dagliati, Mohamad Daniar, Christel Daniel, Priyam Das, Anahita Davoudi, Batsal Devkota, Julien Dubiel, Loic Esteve, Hossein Estiri, Shirley Fan, Robert W Follett, Thomas Ganslandt, Noelia García-Barrio, Lana X Garmire, Nils Gehlenborg, Emily Getzen, Alon Geva, Tobias Gradinger, Alexandre Gramfort, Romain Griffier, Nicolas Griffon, Olivier Grisel, Alba Gutiérrez-Sacristán, David A Hanauer, Christian Haverkamp, Bing He, Darren W Henderson, Martin Hilka, Yuk-Lam Ho, John H Holmes, Chuan Hong, Kenneth M Huling, Meghan R Hutch, Richard W Issitt, Anne Sophie Jannot, Vianney Jouhet, Mark S Keller, Chris J Kennedy, Daniel A Key, Katie Kirchoff, Jeffrey G Klann, Isaac S Kohane, Ian D Krantz, Detlef Kraska, Ashok K Krishnamurthy, Sehi L'Yi, Trang T Le, Judith Leblanc, Guillaume Lemaitre, Leslie Lenert, Damien Leprovost, Molei Liu, Ne Hooi Will Loh, Qi Long, Sara Lozano-Zahonero, Yuan Luo, Kristine E Lynch, Sadiqa Mahmood, Sarah Maidlow, Adeline Makoudjou, Alberto Malovini, Kenneth D Mandl, Chengsheng Mao, Anupama Maram, Patricia Martel, Marcelo R Martins, Jayson S Marwaha, Aaron J Masino, Maria Mazzitelli, Arthur Mensch, Marianna Milano, Marcos F Minicucci, Bertrand Moal, Taha Mohseni Ahooyi, Jason H Moore, Cinta Moraleda, Jeffrey S Morris, Michele Morris, Karyn L Moshal, Sajad Mousavi, Danielle L Mowery, Douglas A Murad, Shawn N Murphy, Thomas P Naughton, Antoine Neuraz, Kee Yuan Ngiam, Wanjiku FM Njoroge, James B Norman, Jihad Obeid, Marina P Okoshi, Karen L Olson, Gilbert S Omenn, Nina Orlova, Brian D Ostasiewski, Nathan P Palmer, Nicolas Paris, Lav P Patel, Miguel Pedrera-Jimenez, Emily R Pfaff, Ashley C Pfaff, Danielle Pillion, Sara Pizzimenti, Hans U Prokosch, Robson A Prudente, Andrea Prunotto, Víctor Quirós-González, Rachel B Ramoni, Maryna Raskin, Siegbert Rieg, Gustavo Roig-Domínguez, Pablo Rojo, Paula Rubio-Mayo, Carlos Sáez, Elisa Salamanca, Malarkodi J Samayamuthu, L. Nelson Sanchez-Pinto, Arnaud Sandrin, Nandhini Santhanam, Janaina CC Santos, Fernando J Sanz Vidorreta, Maria Savino, Emily R Schriver, Petra Schubert, Juergen Schuettler, Luigia Scudeller, Neil J Sebire, Pablo Serrano-Balazote, Patricia Serre, Arnaud Serret-Larmande, Mohsin Shah, Zahra Shakeri, Domenick Silvio, Piotr Sliz, Jiyeon Son, Charles Sonday, Andrew M South, Anastasia Spiridou, Zachary H Strasser, Amelia LM Tan, Bryce WQ Tan, Byorn WL Tan, Suzana E Tanni, Deanne M Taylor, Ana I Terriza-Torres, Valentina Tibollo, Patric Tippmann, Emma MS Toh, Carlo Torti, Enrico M Trecarichi, Yi-Ju Tseng, Andrew K Vallejos, Gael Varoquaux, Margaret E Vella, Guillaume Verdy, Jill-Jênn Vie, Shyam Visweswaran, Michele Vitacca, Kavishwar B Wagholikar, Lemuel R Waitman, Xuan Wang, Demian Wassermann, Griffin M Weber, Martin

Wolkewitz, Scott Wong, Zongqi Xia, Xin Xiong, Ye Ye, Nadir Yehya, William Yuan, Alberto Zambelli, Harrison G Zhang, Daniel Zoeller, Chiara Zucco
